# YouDiagnose Predictive Model Pilot Study to Compare Expert Human vs. Machine Prediction Accuracy

**DOI:** 10.1101/2023.04.18.23288678

**Authors:** Aswini Misro, Vikash Sharma, Naim Kadoglou

**Affiliations:** Innovation Lead, YouDiagnose Limited; Data Scientist, Neurapses Technologies Ltd; Senior Breast Consultant with Interest in Data Informatics, London Northwest University Healthcare NHS Trust

## Abstract

YouDiagnose carried out a pilot study that seeks to compare the accuracy of expert human predictions with those made by a predictive model. Specifically, this study will analyse the prediction of diseases, cancer risk and the care need of patients being admitted or referred to the specialist service. The doctor was provided with a set of patient data, e.g., patient ID 9001 to 9050. The pilot consisted of 2 parts 1. Clinical prediction and 2. Model validation. In the first part, the doctor had to read the information in the clinical vignette and had to select one of the predetermined choices for the best clinical prediction (DP-1, DP-2, DP-3). Afterwards, he/she was asked to validate the model’s predictions (MP-1, MP-2, MP-3) and recommendations (R-1, R-2, R-3) for the same case.

The study shows that MP has higher total accuracy (82.8%) compared to DP (50.6%). In predicting cancer, the MP method has higher sensitivity (100.0%) and positive predictive value (38.5%) compared to the DP method (90.0% sensitivity and 30.0% positive predictive value). The MP method also has higher specificity (78.9%) compared to the DP method (72.7%). Both methods have high negative predictive values (98.2% for DP and 100.0% for MP) with a p-value of 0.3705.

The results of the current pilot study demonstrate the model’s potential, while also highlighting areas where further testing is needed in order to increase user confidence and improve the accuracy of diagnosis. Such testing could provide invaluable insights into how to maximize the value of the system in offering better frontline screening solutions e.g., triaging, clinical decision support, risk-based clinic booking system etc.

## Plain English Summary

YouDiagnose conducted a pilot study comparing the accuracy of predictions made by human experts to the accuracy of prediction models. Specifically, this study examines predictions of disease, cancer risk, and need for care in patients admitted or referred to professional services. A series of patient data were provided to physicians. The pilot projects 2 parts. In the first part, the physician had to read the information in the clinical vignette and write down their best clinical predictions. They were then asked to validate the model’s predictions and recommendations for the same cases.

The study shows higher overall accuracy for the model compared to doctors’ predictions. In cancer prediction, the model was found to have higher sensitivity and positive predictive value. Both methods have a high negative predictive value. The results of the current pilot study demonstrate the potential of the model while highlighting areas where further testing is needed to increase user confidence and improve diagnostic accuracy. Such tests can provide invaluable insight into how to maximize the value of your system by providing better frontline screening solutions. B. Triage, clinical decision support, risk-based clinic appointment systems, etc.

### Ethics approval and consent to participate

At the outset, the research methodology was approved by YouDiagnose Ethical Approval Committee. This research work was commenced after this approval and this study did not involve any non-anonymised human data, tissues, samples, or materials. Consent to participate was obtained from each participant at the beginning of the study. Using the NHS Health Research Authority and United Kingdom Medical Research Council decision-making tool, it was determined that this study would produce generalisable or transferable findings. Therefore, informed consent was obtained from each participant before the structured interview. It was also determined that the study would anonymize the participants to mitigate any risk. All the methodologies were in line with the core practices of the Committee on Publication Ethics (COPE) and the Declaration of Helsinki.

**Figure-1.**
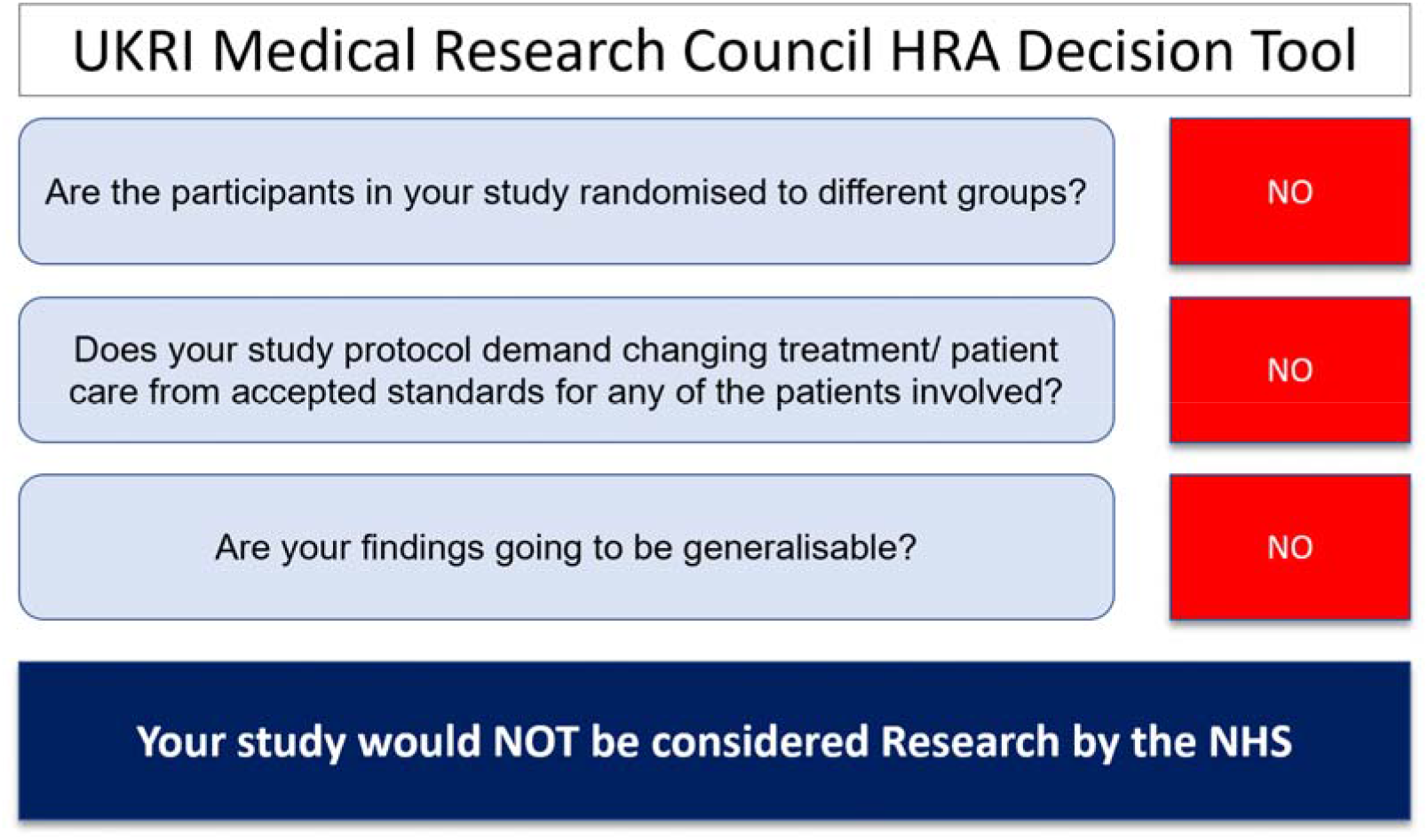
The output of HRA Decision Tool.

## Availability of data and materials

It is our policy to protect the intellectual property of our research and development. Therefore, we cannot share the data which was used to build and test the data model. We understand that this may be a disappointment, but we have shared here an example of YouDiagnose’s Federated Autonomous Multi-Disease Deep Learning Database that was used in 2019 and is now defunct. All rights are reserved by YouDiagnose Limited, and the data must not be used for commercial purposes or distribution. The number associated with the data is the case identifier. This is for reference only and the details cannot be disclosed.

## Source of data

JBS Healthcare, Data partners, & Healthcare organisations

## Background

The nightmare of misdiagnosis has long plagued clinicians, representing the apex of professional adversity. Predictive technology holds great promise to mitigate the risk of erroneous diagnosis or overlooking a crucial aspect of a patient’s condition. Through the utilisation of sophisticated algorithms and data-driven analytics, predictive technology can be leveraged to provide clinicians with invaluable insight into their patients’ conditions, allowing for more accurate, informed, and timely decision-making in the fast-paced clinical environment. This study aims to provide a preliminary proof of concept with secondary healthcare doctors. The purpose of this assessment is to evaluate the accuracy, value, and potential impact of a predictive model that uses data-driven algorithms to process clinical, demographic, and other data. The machine and expert human predictions will be compared by taking the final diagnosis of the diseases as the gold standard. This assessment allows us to gain a better understanding of how these predictive models can help to improve healthcare services. This could be a significant leap forward in improving patient outcomes and boosting healthcare efficiency. We look forward to seeing the results of this important research shaping the future of healthcare decision-making.

### Scientific justification

The use of data-driven risk analytics in the community has tremendous potential to improve the prospect of early prediction of critical diseases e.g., cancer. By providing risk assessment, disease prediction, automation of triaging, workflow automation and management of care pathways, data-driven risk analytics can enable flagging and early diagnosis of life-threatening diseases such as cancer. This could result in improved patient outcomes and better use of healthcare resources. This data-driven risk analytics is an invaluable tool in the fight against serious diseases.

The benefit to patients and the entire ecosystem is clear. Outpatient waiting times can be significantly reduced leading to more accessible healthcare. Additionally, the quality of interactions between patients and their doctors will be greatly improved, with up-to-date and accurate information providing the basis for a more patient-centric user experience. Clinicians too will experience a reduction in professional burn-out due to the lightening of repetitive and administrative tasks. Please see the FAQ section to learn about the use of predictive technology in healthcare.

This pilot study seeks to compare the accuracy of expert human predictions with those made by a predictive model. Specifically, this study will analyse the prediction of diseases, cancer risk and the care needs of patients being admitted or referred to the specialist service. The results from this initial comparison will provide a clearer understanding of the value of such predictive technologies. This pilot study is expected to generate evidence to shape the future of clinical decision-making.

The NHS is facing immense pressure with resource constraints and the backlog of referred cases from the community. Predictive technologies-driven automation can help to improve the system efficiency and cost saving without a huge expense. This technology could enable the NHS to better anticipate demand and manage resources, ensuring that more services can be provided at a much lower cost. With this technological solution, the NHS can rise up to meet the challenges it faces today.

### Principal research question

1. What is the level of agreement between the clinician and the model’s prediction?
2. What is the level of agreement between the doctor’s clinical predictions in comparison to the gold standard e.g., the final diagnosis?
3. What is the level of agreement between the model’s predictions in comparison to the gold standard e.g., the final diagnosis?
4. Please see below in the FAQ section (at the bottom) the meaning of the terms used here e.g., clinician’s prediction and final diagnosis.

### Principal inclusion criteria

The inclusion criteria were as follows.

1. Female patient
2. Over the age of 18
3. Referred by a GP practice/Medical specialities/Self-referrals to hospital services.

### Exclusion criteria

1. Children and youth under the age of 18
2. A male patient
3. Past history of breast cancer
4. Rare cases e.g., Idiopathic Granulomatous Mastitis, Tubercular Mastitis, Mondor’s disease, various rare syndromes etc.

### Sample size

Total sample size:174. We have taken a pragmatic approach and limited the sample size to 174 patients for this proof-of-concept study as it will lay the groundwork for future studies.

### Design and methodology

The study was conducted on the data provided by various healthcare organisations. Out of all the cases, a study sample was selected using the inclusion and exclusion criteria. The selected sample consists of consecutive referrals made from the GP surgery and medical specialities e.g., internal medicine to specialist breast centres. This also includes the cases of self-referrals.

This study will involve the analysis of retrospective data of patients who have been discharged from healthcare practices. The data has been anonymised. Pseudonyms have been used where appropriate. The data has been thoroughly reviewed by breast care professionals to ensure compliance with local standards of practice, and we are aware of some variability. All efforts have been made to ensure that the data presented to participating doctors in the pilot is reliable and accurate.

### Training and induction

The number of doctors included in the study was 9 who saw between 10-43 cases. The doctors were provided with induction on the user interface, software navigation and use of the application. The data-driven application will be tested prior to use by the doctors to ensure there are no software glitches.

### The study and data collection

The study was carried out using Windows or Mac computer laptop connected to the internet. Log-in credentials were provided for each doctor to access the cases. Once logged in, the doctor was asked to click on a case to begin viewing. The timer started automatically and records the time taken for each case. The doctor was provided with a set of patient data, e.g., patient ID 9001 to 9050. The pilot consisted of 2 parts 1. Clinical prediction and 2. Model validation

#### Clinical prediction

Each case contained the clinical details of a patient presented as a story, giving the doctor the opportunity to analyse and make decisions based on their understanding of the situation. The doctor, after reading the information provided, had to select one of the predetermined choices given for each question in order to answer it. Each case had four questions that must be answered to proceed to the next step.

The six questions are as follows.

1. What is your best clinical prediction (for this case)?
2. What is your 2^nd^ best clinical prediction (for this case)?
3. What is your 3rd best clinical prediction (for this case)?
4. What is the cancer risk?
5. What is the urgency of care need?
6. What type of consultation is most appropriate?

#### Model validation

After the prediction section, the clinician was presented with the model’s predictions for the clinical vignette. Here the user (doctor) can choose to agree or disagree with the model results (predictions). This process, called model validation, was designed to verify the level of agreement or concordance between the model and the physician by asking the following questions.

1. Do you agree with Model Prediction-1 (MP-1)?
2. Do you agree with Model Prediction-2 (MP-2)?
3. Do you agree with Model Prediction-3 (MP-3)?
4. Do you agree with Model Recommendation-1 (R-1)?
5. Do you agree with Model Recommendation-2 (R-2)?
6. Do you agree with Model Recommendation-3 (R-3)?

At the end of the assessment, the doctor was asked to provide general feedback on the pilot by rating the performance of the system from 1-10 and giving an evaluation of how well it performed during the assessment. The doctor was also provided with the option to provide any additional comments or insights that could help improve its accuracy and performance in future assessments. The doctor had to log off the system once the data has been submitted.

## Results

The test consisted of 57 types of breast diseases. The dataset contained a total of 174 cases with 20 cases of cancer who had completed their treatment and 154 benign cases. The surgeons spent a total time of 789.46 min, making an average of 4.5 min for each case.

The doctor’s first (DP-1), second (DP-2) and third prediction (DP-3) when matched with the gold standard matched showed an accuracy of 37.9%, 4.6% and 8% respectively. The total number of predictions matching with gold standards across all three predictions was 50.6%. On the other hand, the model’s first (MP-1), second (MP-2) and third prediction (MP-3) when matched with the gold standard matched showed an accuracy of 62.1%, 13.8%, and 6.9% respectively, bringing the total prediction accuracy (e.g., MP-1, 2, & 3) to 82.8%. Please see Table-1.

**Table 1.** Shows a comparison of doctors vs model’s prediction accuracy.

| **Accuracy - Doctor's predictions (DP)** |  |  |  |
| --- | --- | --- | --- |
| **DP-1** | **DP-2** | **DP-3** | **DP-1+2+3** |
| **66** | 8 | 14 | 88 |
| **37.9%** | 4.6% | 8.0% | 50.6% |
| **Accuracy – Model predictions (DP)** |  |  |  |
| **MP-1** | **MP-2** | **MP-3** | **MP-1+2+3** |
| **108** | 24 | 12 | 144 |
| **62.1%** | 13.8% | 6.9% | 82.8% |

If the prediction matched a value of the gold standard, it was called a similarity match. On the other hand, if the prediction matched the gold standard’s exact value, it was called a perfect match. The exact, similarity and total match for DP-1 was 37.9%, 33.3%, and 71.3% respectively whereas the exact, similarity and total match for MP-1 was 62.1%, 23.0%, and 85.1% respectively. Please see Table-2.

**Table 2.** Shows the exact, similarity, and total match doctor vs model’s prediction accuracy.

| **PD-1** | **Doctor's predictions (DP)** | **Model predictions (DP)** |
| --- | --- | --- |
| **Exact match** | 66 | 108 |
| **%** | 37.9% | 62.1% |
| **Similarity match** | 58 | 40 |
| **%** | 33.3% | 23.0% |
| **Total match** | 124 | 148 |
| **%** | 71.3% | 85.1% |

The surgeon (subject matter expert SME) was asked to validate the predictions of the MP (Model Prediction) and they agreed with the prediction of MP-1, MP-2 and MP-3 on 77%, 83.9% and 67.8% respectively. The SME approved MP’s recommendations (R1, R2, R3) more than 85% of the time as can be seen in table-3.

**Table 3.** Shows the doctor’s agreement with model’s predictions and recommendations.

|  | MP-1 | MP-2 | MP-3 |
| --- | --- | --- | --- |
| **Agreement** | 134 | 146 | 118 |
| **%** | 77.0% | 83.9% | 67.8% |
|  | R-1 | R-2 | R-3 |
| **Agreement** | 152 | 150 | 174 |
| **%** | 87.4% | 86.2% | 100.0% |

Table 4&5 shows the sensitivity, specificity, and positive and negative predictive value of predicting cancer using both methods.

**Table 4.**
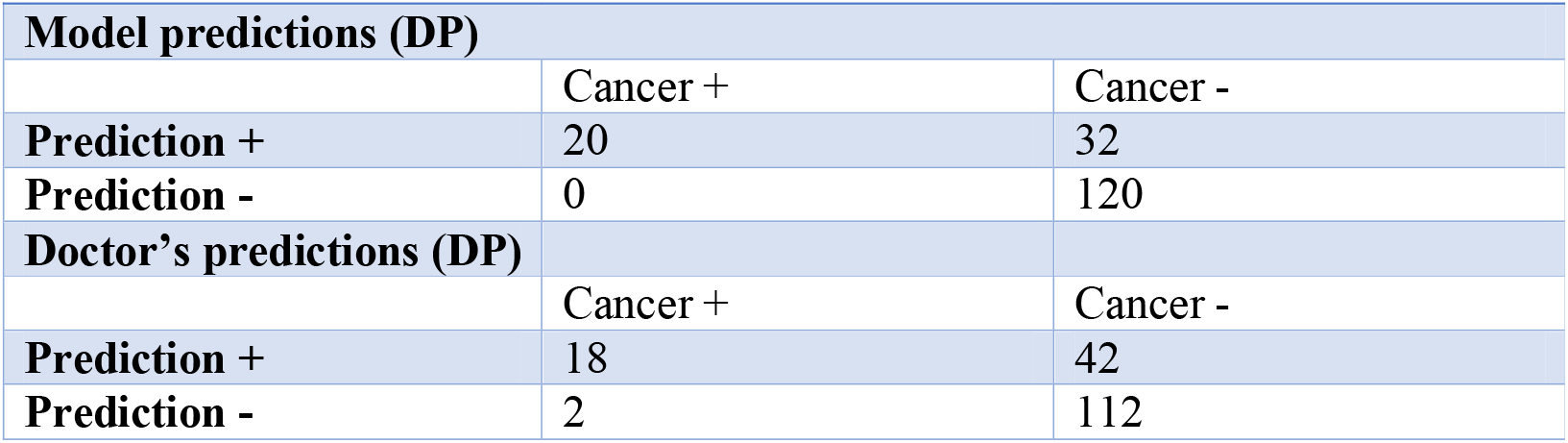
shows the sensitivity, specificity, positive predictive value and negative predictive value for both methods.

**Table 5.** shows the sensitivity, specificity, positive predictive value and negative predictive value for both methods.

| **Parameter** | **DP** | **MP** |
| --- | --- | --- |
| **sensitivity for predicting cancer** | 90% | 100.0% |
| **specificity for predicting cancer** | 72.7% | 78.9% |
| **positive predictive value** | 30.0% | 38.5% |
| **negative predictive value** | 98.2% | 100.0% |

### Observations

Comparing both methods (MP & DP method) using data tables 1-5:

#### Accuracy

The MP has higher total accuracy (82.8%) compared to the DP (50.6%) (Table-1). The MP has a higher percentage of exact matches (62.1%) compared to the DP (37.9%). The MP also has a higher percentage of similarity matches (20.7%) compared to the DP (12.6%). Overall, the MP has a higher total match rate (82.8%) compared to the DP (50.6%) (Table-1).

Sensitivity, specificity, and predictive values in predicting cancer: The MP method has higher sensitivity (100.0%) and positive predictive value (38.5%) compared to the DP method (90.0% sensitivity and 30.0% positive predictive value). The MP method also has higher specificity (78.9%) compared to the DP method (72.7%). Both methods have high negative predictive values (98.2% for DP and 100.0% for MP) (Table-5)

#### SME agreement

The SME agreed with the MP’s recommendations more than 85% of the time, while the agreement rates for the MP were lower (77% for MP-1, 83.9% for MP-2, and 67.8% for MP-3) (Table-3).

Inferring if the difference between both methods is significant: To determine if the differences between both methods are statistically significant, we performed a chi-square test. This test helps to determine if there is a significant difference between the observed frequencies (the data in the tables) and the expected frequencies. For the chi-square test, we used the data from Table-4. After performing the test, if the p-value is less than 0.05, we can conclude that there is a significant difference between the two methods.

Since the p-value (0.3705) from the chi-square test is greater than 0.05, we fail to reject the null hypothesis, which means there is no significant difference between the DP and MP methods in predicting cancers based on the data provided in Table-4. However, this test only compares the methods’ performance in predicting cancers and not other aspects like accuracy, exact matches, and SME agreement. The chi-square test was chosen because it is a widely used statistical method for comparing categorical data, like the data presented in the tables. It helps to determine if there is a significant association between variables, which is applicable to our comparison of the DP and MP methods.

### Limitation of our calculations

Some limitations of the calculations and analysis performed include:

- Sample size: With a sample size of 174, our conclusions may not generalize well to larger populations. Given that there is not enough statistical power to detect small effect sizes. To overcome this limitation, future studies could increase the sample size to improve the power of the analysis. By increasing the sample size, we can enhance the ability to detect significant differences between DP and MP, if they exist.
- Multiple comparisons: We are comparing multiple metrics, which may increase the chance of finding significant differences due to chance alone.
- Lack of information on the nature of predictions: We do not have information about how the predictions were generated or any potential biases in the methods.
- Another limitation is the lack of data on predicted probabilities or scores, which prevents us from comparing the performance of the DP and MP methods using the AUD-ROC. Future studies could collect this data to better evaluate the discriminatory abilities of both methods and identify potential differences in their performance.

To overcome these limitations, a new study with a larger sample size, more information about the prediction methods, and a focus on minimizing multiple comparisons can be conducted. To calculate the size for detecting a significant difference between the 2 methods with a desired power, we need to determine the effect size we want to detect, the significance level (α), and the desired power(1–β). Let us assume that we want to detect a small effect size (Cohen’s h=0.2), with a significance level (α) of 0.05, and a desired power (1-β) of 0.80. In this case, we can use G* power, a statistical software tool for power analysis, to compute the required sample size for McNemar’s test. Using the G*Power, we find that the required sample size for each group is approximately 199. This means that a study with at least 199 samples per group would be necessary to detect a sample effect size with a power of 0.80.

Although our statistical analysis found no significant difference between DP and MP, the later demonstrated some noteworthy advantages. The MP showed higher accuracy for the first prediction (MP-1) at 62.1% compared to DP-1 at 37.9% (Table-1). Additionally, MP achieved a higher exact match rate at 62.1% compared to DP’s 37.9%. Moreover, the Subject Matter Expert (SME) agreed with MP’s recommendations more than 85% of the time, which is a strong indicator of MP’s reliability and practical utility (Table-3) while the MP-1, MP-2, and MP-3 had SME endorsement in 77.0%, 83.9%, and 67.8% of cases (Table-3).

Using the chi-square statistic (0.8026) and the degrees of freedom (1), we can look up the p-value in a chi-square distribution table or use a calculator. The p-value for our test is approximately 0.3705. Since the p-value (0.3705) is greater than 0.05, we fail to reject the null hypothesis, which means there is no significant difference between the DP and MP methods in predicting cancers based on the corrected data provided in Table-4. Keep in mind that this test only compares the methods’ performance in predicting cancers and no other aspects like accuracy, exact matches, and SME agreement.

### Inference

Based on the chi-square test results and the data presented in the tables, we can conclude the following:

- The MP method appears to have higher overall performance in terms of accuracy, exact matches, SME agreement, cancer prediction sensitivity and positive predictive value.
- The DP has a slightly higher specificity, and a higher percentage of similarity matches compared to the MP.

If the chi-square test shows a significant difference between the two methods (p < 0.05), we can conclude that the MP is significantly better than the DP in predicting outcomes. However, if the p-value is not significant, further research with larger sample sizes and additional information about the prediction methods may be needed to make a definitive conclusion.

## Discussion

The mean rate of first correct results for individual SCs ranged between 12% and 61% (Hill, Sim and Mills, 2020). A 2021 review of the symptoms checker showed Twelve tools were identified and included. The mean diagnostic accuracy of the systems was poor, with the correct diagnosis being present in the top five diagnoses in 51.0% (Ceney et al., 2021)

In a 2020 study by Hill, Sim and Mills, the mean rate of first correct results for individual Symptoms Checkers (SCs) ranged between 12% and 61% (Hill, Sim and Mills, 2020). A 2021 review of these tools identified 12 SCs, with an overall mean diagnostic accuracy being poor - the correct diagnosis was only present within the top five diagnoses in 51% of cases (Ceney et al., 2021). These findings suggest that while SCs are relatively accessible to the general public, their efficacy is limited in providing correct medical diagnostics.

Given the poor accuracy of the models, clinicians expressed a lack of confidence in using them for patient care. The reliability and accuracy of these tools are integral to clinician trust; however, most clinicians we interviewed expected the model to be as accurate as a doctor’s prediction (Misro et al., 2023). The poorly designed model without adequate testing when used at the local, regional, or national level results in a very poor impact.

Poorly validated SCs are generally risk-averse. The general public finds it difficult to accept its advice. Having trust issues in face of worrying symptoms can result in poor adoption of symptom checkers (SCs). SCs provide a list of diagnoses at the end of user interaction, however, it is important to note that there are disclaimers to say that these should not be used as a substitute for medical advice in emergencies and are only intended to be used for informational or guidance purposes. Also, SCs caution the user with a warning e.g., “It is possible that a condition might not be suggested here, so it is highly recommended to consult a medical professional if there are any doubts.”

In the community setting, the risks associated with algorithm-based symptom checkers are high. If they face a setback or malfunction, it leads to user rejection and a lack of confidence among users. This in turn causes disinterest in integrating them with the existing electronic patient records and referral systems. The potential benefits of using such systems must be weighed carefully against the risk of user rejection should something go wrong. A cluster-randomised controlled trial and economic evaluation of GP-led triage, nurse-led computer-supported triage, or usual care were conducted to assess the introduction of telephone triage delivered by GPs or nurses. The results showed that this implementation was associated with similar costs to those of the usual care. (Campbell et al., 2014)

The story is more disastrous at the national level. Existing triaging tools used in the country, such as 111, provide an example of a tool that is already in use at the national level and is not making any difference despite the huge expense of £33 million a year (Burgess, n.d.). The results of the study indicate that the implementation of the NHS 111 online service had a negligible effect on the number of calls to NHS 111 and total triaged calls (Simpson et al., 2022). This suggests that the workload for the telephone service has not changed because of the online service. Furthermore, there is evidence to suggest an increase in referrals to emergency departments, general practitioners, and ambulances since the introduction of NHS 111 online. Altogether, it appears that while NHS 111 online may have had some negative impact on referrals to other services, it has not reduced or increased calls to NHS 111 itself.

In conclusion, it is clear that more research is necessary to improve the accuracy of medical algorithms. The current studies into the 111 system demonstrate that it functions as an issue-based segregation tool rather than a disease prediction system. This lack of accuracy compromises stakeholders’ confidence in using such systems and thus necessitates a dedicated effort to increase their reliability.

The current proposed YouDiagnose model has been designed with the goal of meeting both doctors’ and patients’ need. YouDiagnose has already a usability evaluation was conducted with participants from the Patient and Public Involvement and Engagement Senate (PIES) of the Innovation Agency (an Academic Health Science Network) Qualitative feedback was obtained from the participants on both modalities and quantitative feedback in the form of the System Usability Scale (SUS), comparing the usability of both interaction modalities. The SUS scores were analysed using the Adjective Rating Scale that revealed the Smart Questionnaire had ‘Good Usability’ compared to ‘OK Usability’ of the Chatbot. (Misro et al., 2022).

The results of the current pilot study demonstrate the model’s potential, while also highlighting areas where further testing is needed in order to increase user confidence and improve the accuracy of diagnosis. Such testing could provide invaluable insights into how to maximize the value of the system in offering better diagnostic solutions.

## Data Availability

All data produced in the present study are available upon reasonable request to the authors

## Glossary of terms

1. Similarity match: ‘Similarity match’ term is used when there was a match with a disease which is similar in etiopathogenesis, sharing the presenting features and at least the initial line of treatment. For example, certain types of diseases cannot be distinguished from one another based on presentation, and it is almost impossible to distinguish between them based on medical history alone. Needs scrutiny. However, there are many diseases that share similar aetiology, pathogenesis, symptoms, initial plan, and treatment. Examples can be given of lactation mastitis and breast abscess which are infections involving breast tissue in a breastfeeding woman and need prompt treatment with antibiotics. Both have the same symptoms and cannot be distinguished without ultrasound.
2. Gold standard: For all cases, the final diagnosis or final diagnosis was accepted as the disease the patient had, as confirmed from the patient’s case record after the completion of investigations and interventions. This was taken as the gold standard for comparing model performance.

## Addendum-1 – How p-value was calculated?

Table-4 (Recalculated)

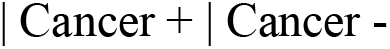
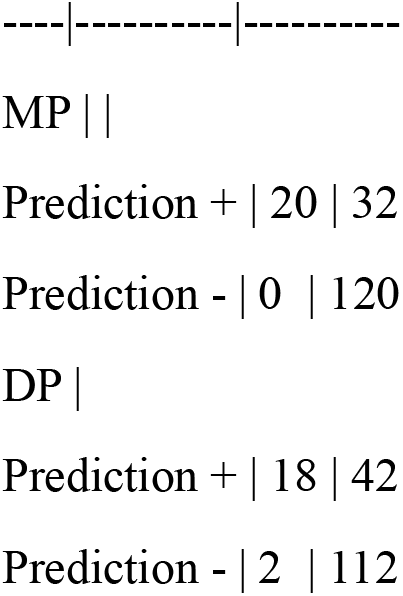

To perform the chi-square test, we first create a contingency table:

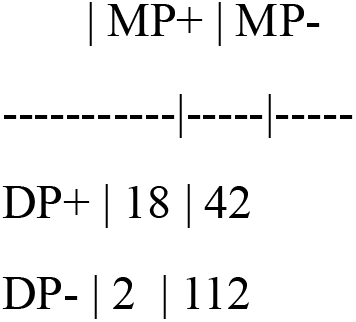

We then calculate the expected values for each cell in the contingency table under the assumption that there is no association between the DP and MP methods. To do this, we use the following formula for each cell: (row total * column total) / grand total.

Expected values:

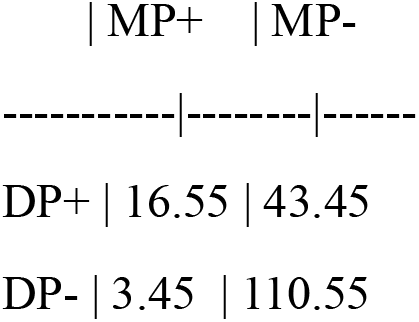

Now, we calculate the chi-square statistic using the formula: X^2^ = Σ[(observed - expected)2 / expected]

X^2^ = ((18 - 16.55)^2^ / 16.55) + ((42 - 43.45)^2^ / 43.45) + ((2 - 3.45)^2^ / 3.45) + ((112 - 110.55)^2^ / 110.55) = 0.1277 + 0.0478 + 0.6082 + 0.0189 = 0.8026

Next, we find the degrees of freedom (df) for the test. In this case, df = (number of rows - 1) * (number of columns - 1) = 1 * 1 = 1.

Using the chi-square statistic (0.8026) and the degrees of freedom (1), we can look up the p-value in a chi-square distribution table or use a calculator. The p-value for our test is approximately 0.3705.

Since the p-value (0.3705) is greater than 0.05, we fail to reject the null hypothesis, which means there is no significant difference between the DP and MP methods in predicting cancers based on the corrected data provided in Table-4. Keep in mind that this test only compares the methods’ performance in predicting cancers and not other aspects like accuracy, exact matches, and SME agreement.

